# Evaluation of a participatory approach to stratifying neighbourhoods across the Tanzanian city of Dar es Salaam in terms of income by comparing with latent profile analysis of deprivation based on national census data

**DOI:** 10.1101/2024.04.19.24305791

**Authors:** Ibrahim Msuya, Martin Boudou, Francis Levira, Irene Moshi, Jean O’Dwyer, Gerard Killeen

## Abstract

Studying geographic heterogeneities in the characteristics of city neighbourhoods, such as population income and deprivation, has been a common practice in urban health studies for tailored and targeted interventions, especially in highly developed and diversified cities. While a range of different approaches has been applied to classify different parts of towns and cities, participatory stratification approaches (PSAs) have become popular despite their subjective basis because of their affordability, simplicity and practicality, all of which allow them to be frequently updated. While more objective statistical approaches, such as latent profile analysis (LPA), can also be used to stratify neighbourhoods using formal socio-economic and demographic data, these rely on the availability of rich datasets and advanced analytical capacities that are not always available in low and middle-income countries. This study assessed a PSA to stratify neighbourhoods across the Tanzania city of Dar es Salaam in terms of income, by comparing it with a complementary LPA using national census data from 2012 to stratify them in terms of deprivation. A consultative community-based workshop was used for the PSA, while 15 selected deprivation indicators from the census data were used to profile them using LPA. While the PSA allocated neighbourhoods to five income strata, six clear deprivation strata could be distinguished by LPA. A strong positive correlation was observed between the stratum identified by the LPA and that obtained through the PSA (ρ = 0.739, p < 0.0001). Furthermore, paired comparison of the two sets of correlation coefficients between each deprivation indicator and the stratum assigned by each stratification approach revealed no difference (V = 33, p-value = 0.1354), confirming that the two approaches yielded very similar patterns of stratification., Also, the two approaches yielded broadly comparable cartographic pictures of the city, depicting similar spatial distribution of wealth and poverty. Overall, this evidence indicates that subjective community knowledge and lived experience may be invaluable for understanding the built environment and for mapping out pockets of poverty and affluence at fine scales with minimum resources.

## Introduction

More than half of the world’s population now lives in towns and cities [1]. While Africa and eastern and southern Asia have been urbanized most rapidly, the pace of urbanization in these regions is expected to slow over the coming decade [2]. Meanwhile, limited government investment in urban planning has contributed to more extensive and overcrowded informal settlements, where approximately 47% of the world’s 4.3 billion urban dwellers live [2]. In Tanzania, for instance, it has been estimated that more than 75% of settlements have been informally developed [3]. Indeed, Dar es Salaam is the largest city in Tanzania, where >75% of the population live in informal settlements with highly heterogeneous wealth distribution [4]. Several historical studies on the early development of what is now the old city centre of Dar es Salaam have rationalized the distribution of settlement patterns in terms of colonial planning processes, which were heavily influenced by residential segregation policies. During the colonial era, the city was planned in a way that segregated people by race and income, with affluent neighbourhoods located near the city centre and along the Indian Ocean (Europeans and Indians) and the less affluent near and around the city centre (black Africans, referred to in that era as natives) (Brennan & Burton, 2007). The evident pattern in the city demonstrates that a significant portion of its development has unfolded through informal processes [5]. Informal settlements tend to be situated either within formal areas or near them, aligning with the contextual reality of the city’s historical evolution. This historical development trajectory has contributed to an uneven distribution of neighbourhood attributes in terms of income and affluence, a disparity that persists today. However, despite the inclination to attribute these disparities solely to colonial influences, a more thorough investigation challenges this prevailing narrative. Contrary to the common assumption that the current state of Dar es Salaam, beyond the old city, is a direct result of colonial planning [6], in fact, it emerged from limited planning, especially during the city’s rapid expansion from the 1970’s onward [7]

While both formal and informal settlements co-exist side-by-side in cities, they exhibit varying neighbourhood characteristics. Urban neighbourhoods constantly evolve at very fine scales, resulting in a myriad of spatial, sociodemographic, economic, and cultural diversities coexisting within relatively small, but nevertheless, very heterogeneous geographic areas. This vibrant evolutionary process is particularly dynamic in unplanned, informal settlements, catalyzing different types of disparities, such as health inequality, social division, exclusion, segregation, which are typically assessed in terms of income inequality or socioeconomic deprivation, [8]. Despite the conceptual differences in how income and deprivation are used to assess the two-sided coin of poverty and wealth (Box 1), either may be just as readily applied to surveying the distribution of inequalities across urban areas.

### Box 1.

**Summary of the conceptual and methodological distinctions between *income inequality* and *socioeconomic deprivation*.**

**Income inequality:** The concept of *income inequality* specifically targets the unequal allocation of income among individuals or households within a given population. Surveyed indices of income inequality express the disparity between the earnings of the wealthiest and the poorest members of society in semi-quantitative terms, often employing statistical summary measures, such as the Gini coefficient or income quintiles [9].

**Socioeconomic deprivation:** On the other hand, *socioeconomic deprivation* refers to disadvantages that a person or group face in an inequitable manner [10]. Such disadvantages can be linked to geographic factors, such as living in a certain neighbourhood or community, or other factors like housing and built environment quality. These disadvantages can take various forms, such as limited access to job opportunities, education, healthcare, and other basic needs. Essentially, it describes the relative absence of power that certain individuals or groups have over their economic and social circumstances [11]. This can perpetuate a cycle of poverty, marginalization, and exclusion, leading to increased social inequality and decreased opportunities for upward mobility [12]. Semi-quantitative measures of socioeconomic deprivation often involves use multidimensional indices to account for these diverse factors affecting well-being [9].

For the first time within the context of internationally agreed development goals, the 2030 Agenda aims to reduce inequality based on income [13]. While efforts are being made to bridge the gap between rich and poor [14, 15], there is still insufficient investment in methodologies for socioeconomic profiling of urban neighbourhoods [16]. In response to that methodological need, this study set out to evaluate a subjective participatory stratification approach (PSA) to classifying neighbourhoods across the Tanzanian city of Dar es Salaam in terms of income, by comparing it with an objective latent profile analysis (LPA) of deprivation based on national census data (Box 2).

### Box 2.

**Summary of the relevant evidence base and methodological considerations that informed the formulation of the study design.**

**Participatory Stratification Approaches (PSA):** Participatory stratification approaches involve active, collaborative engagement and involvement of participants who are residents of the study area in question [17, 18]. These participants may sometimes be influential or authoritative stakeholders, in the research investigation or decision-making process. PSA can be used to understand socioeconomic, geographical inequalities and the hierarchical arrangement of social groups within a given society or system, and is especially useful where socio-economic classes are distributed very unevenly among people within a population [19, 20], forcing people with comparable socio-economic characteristics to reside in similar spatial localities [21]. However, it is essential to recognise the subjective nature of PSA, which can be influenced by relative income assessments within the social circles or communities of individual participants, drawing from lived experiences and personal knowledge [22–24]. Nonetheless, such subjective impressions of individual members within a community can yield qualitative insights that complement objective quantitative measurements, thus enriching an understanding of the broader economic landscape within cities [25, 26].

**Latent Profile Analysis (LPA):** A more objective way to understand such socioeconomic strata within in urban spatial and administrative subunits, is through statistical analysis of more structured empirical data from surveys of the major drivers and consequences of such heterogeneities. Specifically, LPA is a statistical method used to identify unobserved subgroups, or latent class or profiles within a sample of observed entities, based on similar patterns observed variables [27]. LPA has been sometimes referred to as an advanced model-based clustering approach, due to its ability to use different fit statistics to select the best profile, class, or cluster. Over the last decade, LPA has become more widely applied as an alternative to previously existing methods for identifying clusters (eg. K-means clustering, Hierarchical clustering, Fuzzy C-means clustering, and Partitioning Around Medoids), due its flexibility in model criterion selection, such as variance and covariance estimation, and the objectivity of its best fit model selection procedures [28–31]. LPA of detailed data from the 2012 national Population and Housing Census (PHC) for Tanzania was therefore adopted as the gold standard objective statistical methodology, with which the PSA was compared and contrasted as a means to assess the advantages and disadvantages of the latter relative to the former.

## Methods

### Ethics statement

This research was conducted as part of Centre for Sustainable, Healthy, and Learning Cities and Neighbourhoods, supported by funding from UK Research and Innovation (UKRI) through the Economic and Social Research Council (ESRC) under the UK Government’s Global Challenges Research Fund (GCRF) with project reference (ES/P011020/1). Ethical clearance for the study, a component of the Sustainable Cities project at Ifakara Health Institute (IHI), was granted by the Institutional Review Board of the Ifakara Health Institute (IHI/IRB/NO: 26-2018) and the Medical Research Coordinating Committee at the National Institute for Medical Research in Tanzania (NIMR/HQ/R.8a/Vol. IX/2954). Verbal consent, in accordance with the approved protocols, was obtained from all human participants.

#### Request for Access to Restricted Materials

Certain materials, such as photographs featuring individuals or identifying body parts, have been omitted from this publication (e.g., Figure 2, panels A, C, D, E, F) in accordance with editorial guidelines. We recognise the significance of these materials to your understanding of the study. Please contact the corresponding author for assistance if you require access to these materials.

### Geographical context and study data

Dar es Salaam is the largest city in Tanzania, located on the Indian Ocean coast (Fig 1). The city had a population of approximately 4.4 million in 2012 but currently has approximately 5.4 million inhabitants, distributed across an administrative area that has remained constant at 1,590 km^2^ [32]. It is also the port in Tanzania, contributing substantially to the national economy and served as a government administrative centre before *de facto* relocation to the official capital, Dodoma, was practically implemented in 2016 [33].

**Fig 1:**
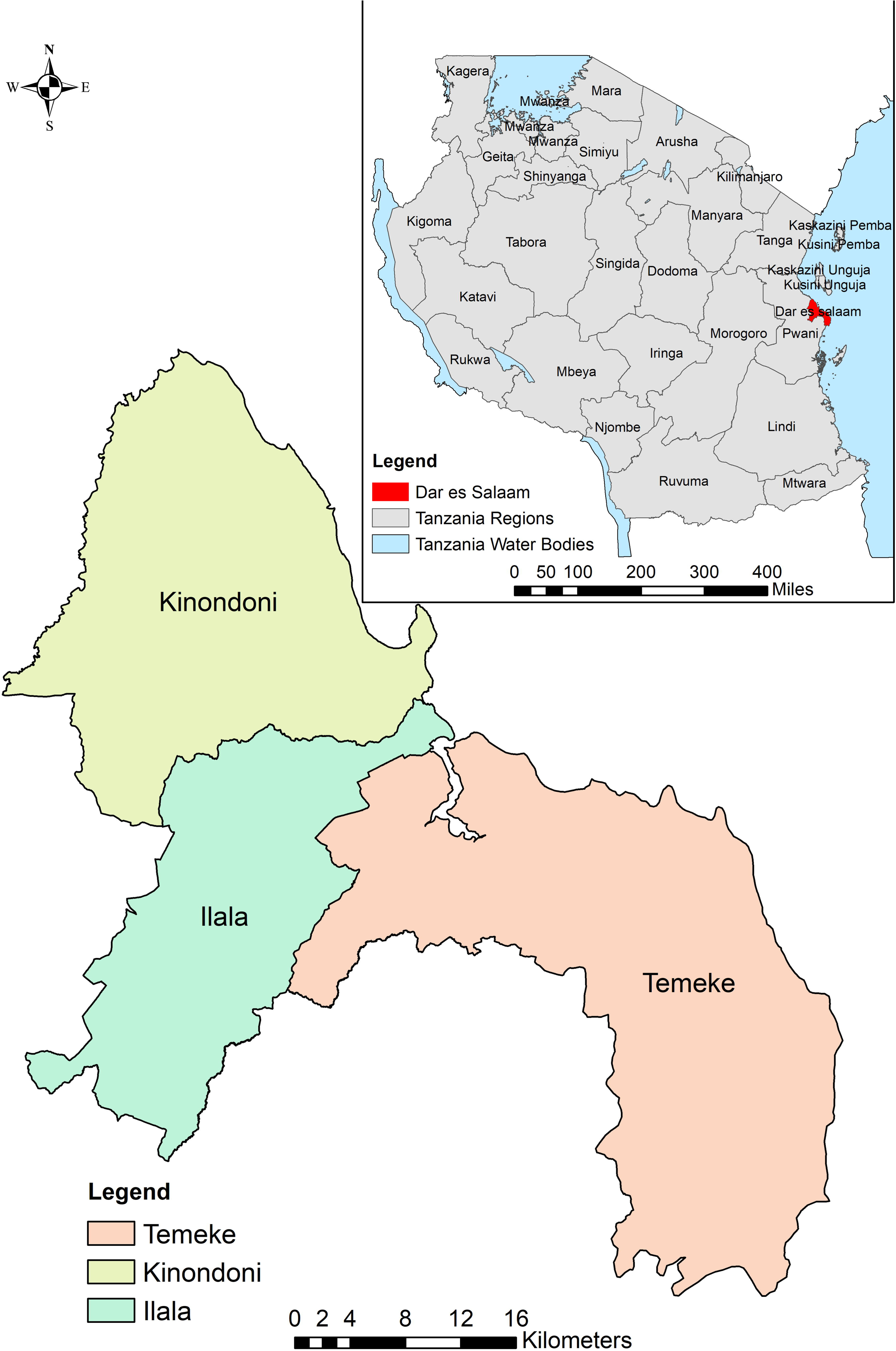
National and regional setting of Dar es Salaam city with district’s boundaries in 2012 (A), wards boundaries (B), and neighbourhood boundaries, the finest administrative scale which could be accessed and used in the study (C).

The study reported herein used Tanzania’s 2012 Population and Housing Census (PHC) data from the National Bureau of Statistics of Tanzania (NBS) to generate variables which were considered likely to represent deprivation in the study context (S1 Table). The data is presented via administrative units, starting from regions down to districts, wards (*kata*) and neighbourhoods (*mitaa*, literally translated as *streets* but actually means a larger, formalized administrative area with defined boundaries, rather than just a road and structures built along it). The data from PHC primarily consisted of socio-demographic information such as gender, age, disability, marital status, citizenship, place of residence, education status, migration patterns, economic activities, fertility rates, general and maternal mortality rates, and enrolment in social security programs. Other data include household with members who are living in diaspora, food and cash crops cultivation, and information about asset ownership and housing conditions. The current study used data aggregated at the neighbourhood level (Fig 1C), the finest scale freely available to the public upon request for research purposes to derive deprivation indicators based relevant variables in the PHC (S1 Table). The dataset included 452 neighbourhoods from three districts that comprised Dar es Salaam at the time (Fig 1A)

Neighbourhoods in Dar es Salaam city are unevenly distributed, with Kinondoni District having the highest number of neighbourhoods, while Ilala has the lowest number of neighbourhoods (Table 1). In terms of population, Kinondoni also has the highest share of residents living in this district, while Ilala is the least. However, from a historical perspective, Ilala stands as the most well-established district, comprising neighbourhoods with a rich legacy that dates back over a century. On the other hand, most parts of Temeke were relatively new and were experiencing rapid growth and development at the time.

**Table 1:** Population and neighbourhoods across districts in Dar es Salaam.

| District | No. Neighbourhoods | % Neighbourhoods | Population | % Population | Area (sq. km) | Population Density (people/area sq. km) |
| --- | --- | --- | --- | --- | --- | --- |
| Kinondoni | 171 | 37.83 | 1,775,049 | 40.67 | 512.71 | 3462.07 |
| Ilala | 101 | 22.35 | 1,220,611 | 27.97 | 389.63 | 3132.72 |
| Temeke | 180 | 39.82 | 1,368,881 | 31.36 | 728.65 | 1878.64 |
| **Total** | **452** | **100.00** | **4,364,541** | **100.00** |  |  |

### Study design

Based on considerations of the relevant methodological literature (Box 2), the current study sought to assess a subjective participatory stratification approach (PSA) to classifying neighbourhoods in terms of income level by comparing it with an objective statistical approach using latent profile analysis (LPA), to stratify neighbourhoods in terms of deprivation based on socioeconomic data from the 2012 PHC in the Tanzania city of Dar es Salaam. While the income which is referred by the PSA is inherently subjective and based on the lived experiences and knowledge of the city’s residents, the LPA of deprivation strata in quantitative terms is a more objective socioeconomic metric derived from formal ways of living standards, conditions, and education at the neighbourhood level [34].

#### Participatory stratification approach (PSA)

The PSA involved active participation of community members and professionals in the stratification of Dar es Salaam neighbourhoods into different income strata. This one-day workshop, carried out on Friday, 22^nd^ March 2019, from 09:00 AM to 17:00 PM at Ifakara Health Institute offices in Mikocheni, Dar es Salaam, involved 50 people, specifically five residents from each district recommended by the neighbourhood leaders, two town planners from each district, three community development officers from each district (Table 1), five practicing town planners from the private sector, two urban and regional planning lecturers from Ardhi University and 13 Ifakara Health Institute scientists working in urban projects in Dar es Salaam. The task started by collectively agreeing on the number of income strata to which the city’s neighbourhoods should be assigned. Then, each of the 452 neighbourhoods were assigned to a respective stratum by each individual participant, following which a consensus was established through group discussion to collectively agree on the stratum it was finally assigned to (Fig 2). Although the task fundamentally relied on the participants knowledge and lived experience of the city, some other complementary sources of information, such as satellite images, were also used to facilitate the process of informed debate and consensus building.

**Fig 2:**
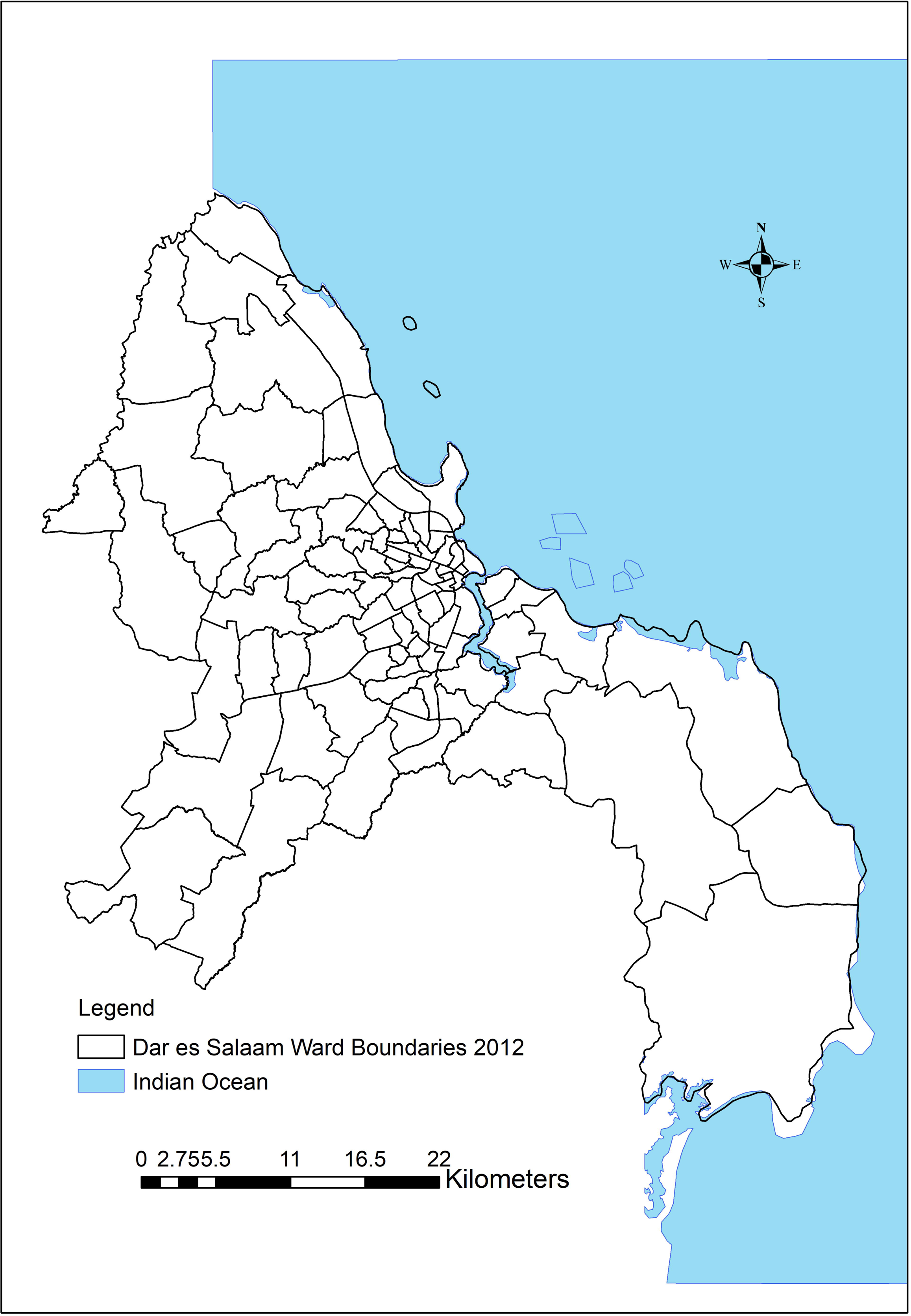
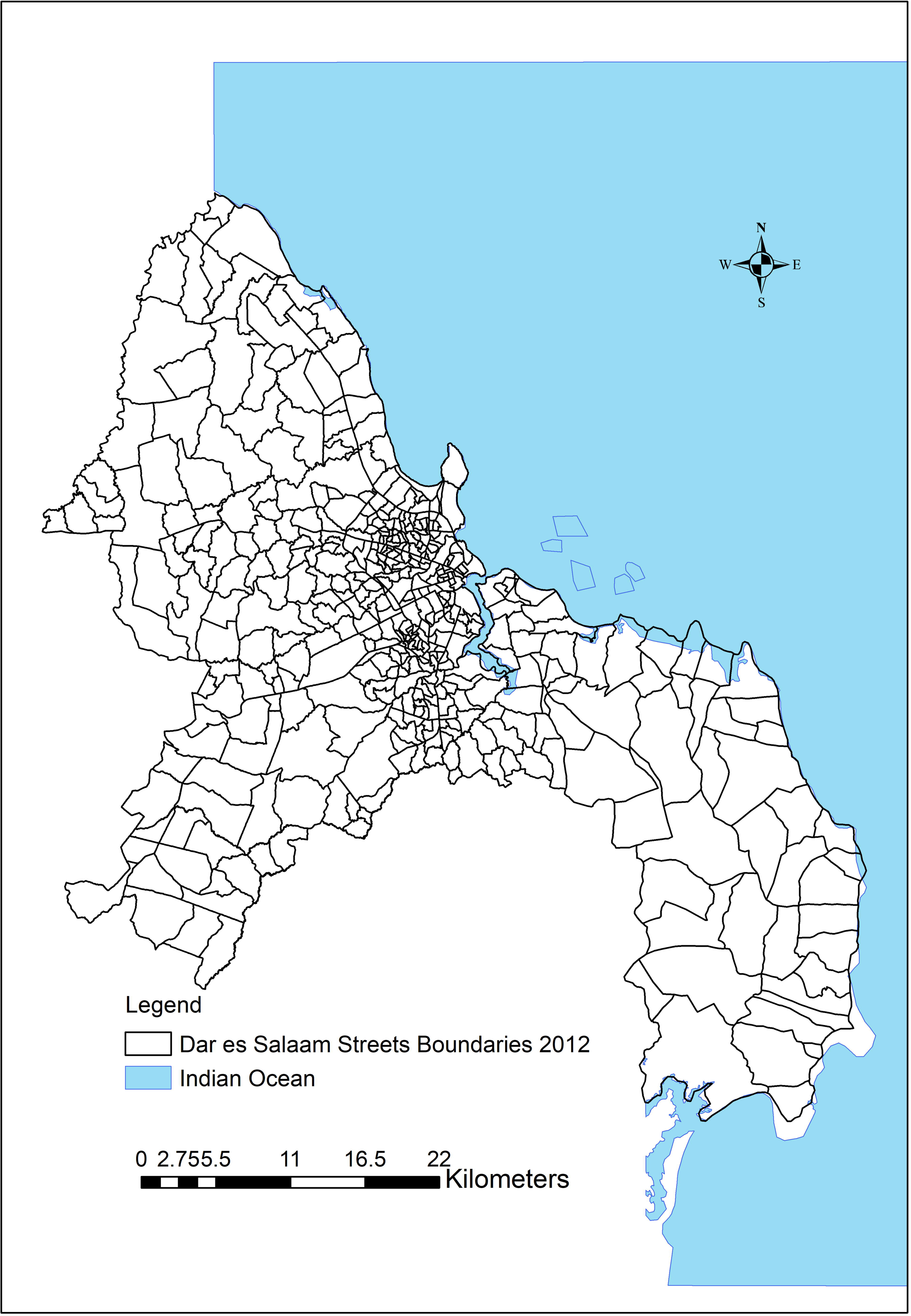
Images showing participants and activities conducted during the participatory neighbourhood stratification workshop. Panel A and B show individual participants independently assigning neighbourhoods to one of five different income strata and sharing their assignation using sticky pad notes. Panels C and D show how the assignations of individual participants were collated and compared as basis for group discussion, by assembling such sticky notes on a flip chart. Panels E and F illustrate how the investigators then facilitated collective consensus building among the participants, based on group discussions to compare and contrast their different neighbourhood assignments to specific strata, and then reach final agreement for each neighbourhood.

#### Latent profile analysis (LPA)

Deprivation profiles were identified as unobserved variables (latent profiles), which were distinguished based on the observed values for relevant socioeconomic variables in the neighbourhood-level PHC data. The first step of the LPA was the identification of the most appropriate recorded variables. Based on the data and context, 15 variables in the PHC data that were intuitively to expected to be closely associated with deprivation were identified. These included education indicators such as; *proportion completed any school level, proportion educated to university level, proportion educated to secondary level,* literacy and language proficiency; *proportion of literate, proportion of English literacy,* economic factors; *proportion of potentially economically active individuals, proportion enrolled in social security, proportion household with members in diaspora,* social and legal indicators*; proportion with registered birth certificate*, demographic factors; *average household size, proportion of female, proportion of single head households,* and health and mortality statistics; *proportion with disability, proportion of death caused by diseases,* and *crude death rate.* Different indicators from the census data were used to derive these variables (S1 Table) for participating respondents or households, aggregated at neighbourhood level. To provide a standardised way to compare different quantities while allowing for more accurate and meaningful comparisons between variables in predicting outcomes or estimate probabilities, almost all variables were transformed into proportions, except for household size and crude death rate which were presented in their actual metrics units.

Descriptive statistics of the selected variables can be found in Fig 3, which shows prior expected general distribution of income and poverty across three study districts at the time, which are consistent with the actual existing situation in the city.

**Fig 3:**
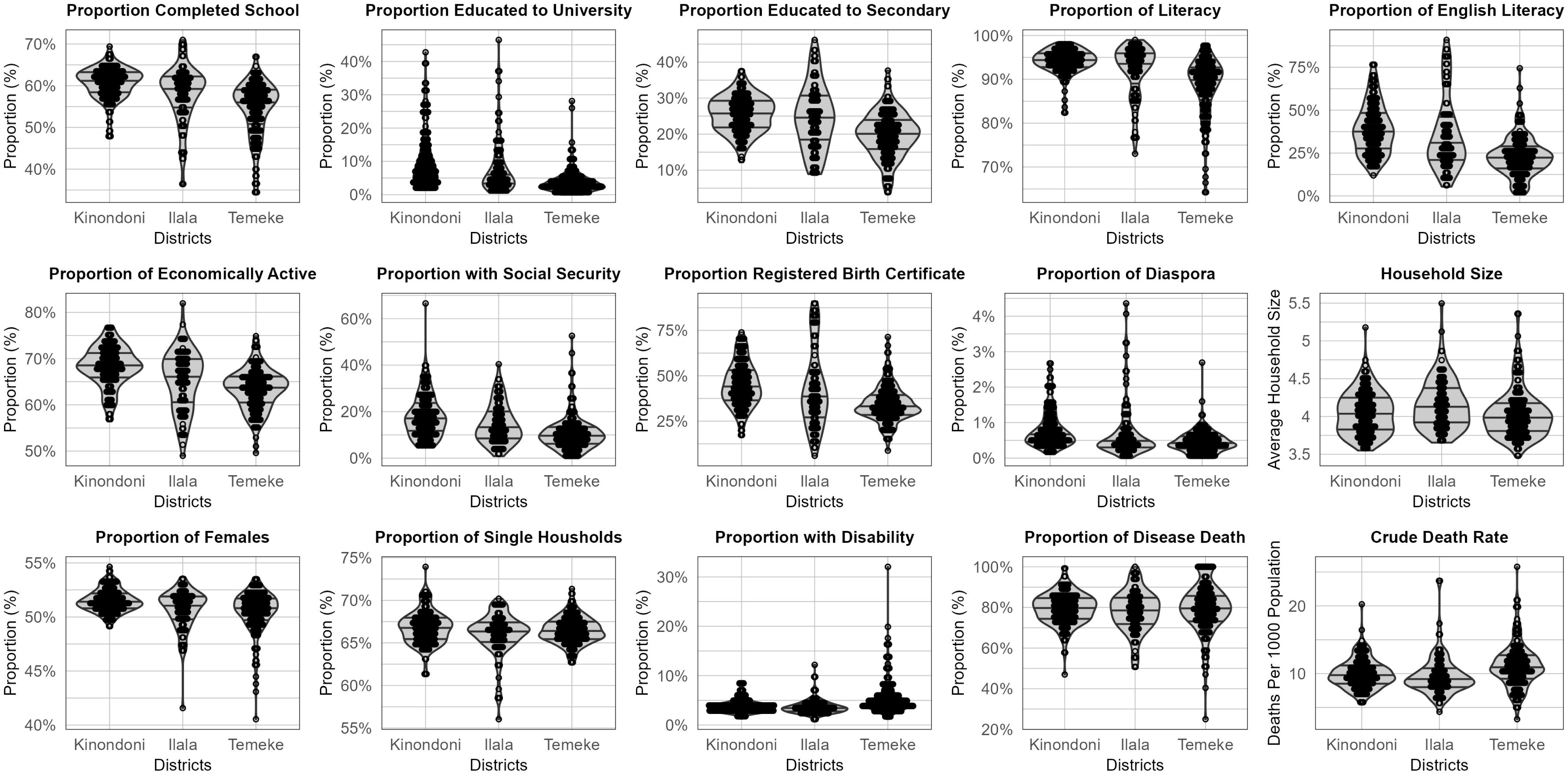
Violin and dot plot showing the descriptive statistics of 15 potential indicators of deprivation derived from the 2012 national census data, distributed across three study districts at the time. Horizontal lines within the violin plots represent the first, second and third quartiles, or the 25%, 50% (median), and 75% quantiles. The units for most of these variables are proportions of participating respondents or households, aggregated at neighbourhood level, the only two exceptions being household size and crude death rate.

Using the selected 15 variables from the neighbourhoods, LPA was used to profile neighbourhoods into statistically distinct strata.

Depending on the nature of data, different six LPA models structures can be used which range from simple to complex, determined by how variance and covariance are treated and whether relationships between variables are considered, thus affecting model parsimony and complexity [35]. In model selection process, Pearson’s rank correlation test was employed to quantify the level of correlation to inform optional specification of model assumption and maximize model performance. In addition, the correlations between variables can help with the interpretation of the resulting latent profiles, as such, if two variables exhibit high level of correlation it can suggest that these two variables may be driving the occurrence of profile and provide insights into the nature of the profile. Broadly speaking, a correlation coefficient ρ ≥ +/- 0.4 and p value ≤ 0.05 was considered as an indication of strong correlation [36]. The Pearson’s rank correlation analysis revealed that a significant correlation existed among ten out of the fifteen variables (p ≤ 0.05), (S2 Fig and S3 Fig). Hence, from the six tidyLPA models structures proposed by Rosenberg *et al.* [29], model structure six with varying variances and covariances was selected. It is the most complex model, so while it allows for the estimation of variances and covariances across profiles without constraints, potentially providing a deeper understanding of the relationships between the variables, it is also less parsimonious.

The LPA modelling was performed using R version 4.2.0 and RStudio version 2023.03.0+386 with the *tidyLPA* package. *tidyLPA* acts as an interface to the *mclust* package and provides the functionalities to estimate the commonly specified models used in the analysis, with different parameters depending on model complexity according to the structural forms developed by [35]. Seven criteria were used for best fit model selection: Log-Likelihood (LogLik), Akaike Information Criterion (AIC), Difference in AIC values (ΔAIC), Consistent Akaike Information Criterion (CLC), Kullback-Leibler Information Criterion (KIC), Sample-Size Adjusted Bayesian Information Criterion (SABIC), and Entropy (Average Uncertainty or Randomness in Latent Class Assignments). Across the existing LPA literature, it has been widely reported that selection of the best fit model with the most informative number of distinct profiles remains a challenging task [37]. Correspondingly, to validate the model optimization based on the fit statistics, an analytic hierarchy process (AHP), as proposed by [38] which helps in determining the correct number of profiles in a model based clustering was used.

Two approaches were used to interpret and assign meaning to each profile from the selected best fit model in relation to deprivation. Firstly, the distribution of the variables was plotted against every profile in a dot and violin plot. Secondly, using z-score or scaled mean of each variable on each profile. While median centre data around the second quartiles or 50% quantiles, z score means centre data around zero because the scaling involves standardizing each observed variable to have mean of zero and standard deviation of one (μ = 0, σ = 1), [39]. In the context of LPA, scaled means are often reported as part of the model output along with other fit statistics. Often, scaled means may have negative value due to scaling even though latent variable has only positive values, however, this does not necessarily indicate that the underlying construct is negative [40]. This helps in comparing the relative positions of the means across variables with different scales or variances, as well as for interpreting the magnitude and direction of the differences between the means of different profiles.

In LPA, after numbering profiles by ranking them according to their order of score, it is always a task to the investigators to interpret and assign their intuitive meaning [41]. The ranked deprivation profiles were then each interpreted and assigned to named ranked deprivation stratum. The LPA deprivation strata were then compared to the results from the PSA using a non-parametric statistical correlation test (Spearman’s rank correlation test), to assess the strength and direction of the relationship between the ranked strata from the two approaches. Further analyses were performed to cross validate the PSA income strata by plotting values using violin and dot plot for the same PHC variables used in broken down by PSA stratum. To test statistical significance difference between the two approaches, a Wilcoxon signed-rank test, was applied using to compare the correlation coefficient of the Spearman’s correlation (ρ) obtained by PSA and LPA.

Additionally, using the ranked results from the two approaches, neighbourhood income and deprivation strata from PSA and LPA, respectively, were mapped using ArcGIS Pro 3.1.2, to understand and compare stratification of the city neighbourhoods by the two approaches in terms of comparative spatial distribution. In spatial comparative analysis, ground truthing evidence of the existing conditions in the selected neighbourhoods, specifically, housing density and quality of buildings, using in situ ground photographs and satellite images, were used to provide further descriptive insights into the meaning of the results.

## Results

### Latent profile identification

The best fit profile model identified by LPA of the census data was with 6-profiles: The goodness of fit consistently decreased as the number of profiles modelled increased, while entropy increased with a 98% classification quality reached for this final optimal 6-profile model that was the last one to converge (Table 2). These model optimization results were confirmed using the AHP approach [38], which also indicated that the model with 6 profiles was optimal based on the fit statistics in Table 2. The results, therefore, suggested that six clear deprivation strata could be identified based on census data, as compared to the five income strata obtained from the participatory stratification approach.

**Table 2:** Comparison of model fit statistics from different models with different numbers of profiles using the tidyLPA complex model structure 6 with varying variance and covariance [29], which were then evaluated using an Analytic Hierarchy Process (AHP) [38] in the selection of the best fit model.

| No. Profile in a Model | LogLik | AIC | ΔAIC | CLC | KIC | SABIC | Entropy |
| --- | --- | --- | --- | --- | --- | --- | --- |
| 1 | -6128 | 12527 | N/A | 12259 | 12665 | 12654 | 1 |
| 2 | -4738 | 10017 | 2510 | 9477 | 10291 | 10272 | 0.96 |
| 3 | -4486 | 9786 | 231 | 8974 | 10196 | 10169 | 0.95 |
| 4 | -3945 | 8977 | 809 | 7893 | 9523 | 9487 | 0.97 |
| 5 | -3677 | 8712 | 265 | 7356 | 9394 | 9350 | 0.98 |
| **6** | **-3435** | **8500** | **212** | **6872** | **9318** | **9267** | **0.98** |

### Interpreting the underlying meaning of identified latent profiles

From the dot and violin plots in Fig, the value distributions of the most important individual indicators of deprivation show that ranking the numbered LPA profiles based on these same variables, yields an order of 2, 1, 3, 4, 5 and 6 for the profile numbers assigned by the LPA best fit model (Table 2). Correspondingly, these were respectively assigned the following stratum names; *Highly Deprived, Deprived, Moderately Deprived, Moderately Affluent, Affluent,* and *Highly Affluent* (Table 3). To further examine and query these profile assignments based on variable value distributions, a calculated scaled mean for each variable was used (Fig 5), consistent with other LPA studies that assigned interpreted profile meanings [28, 42–44]. These scaled means for each variable within each stratum confirmed the same apparent profile order (Table 3) for most variables, as ranked from the most deprived to the most affluent (Fig 5).

**Fig 4:**
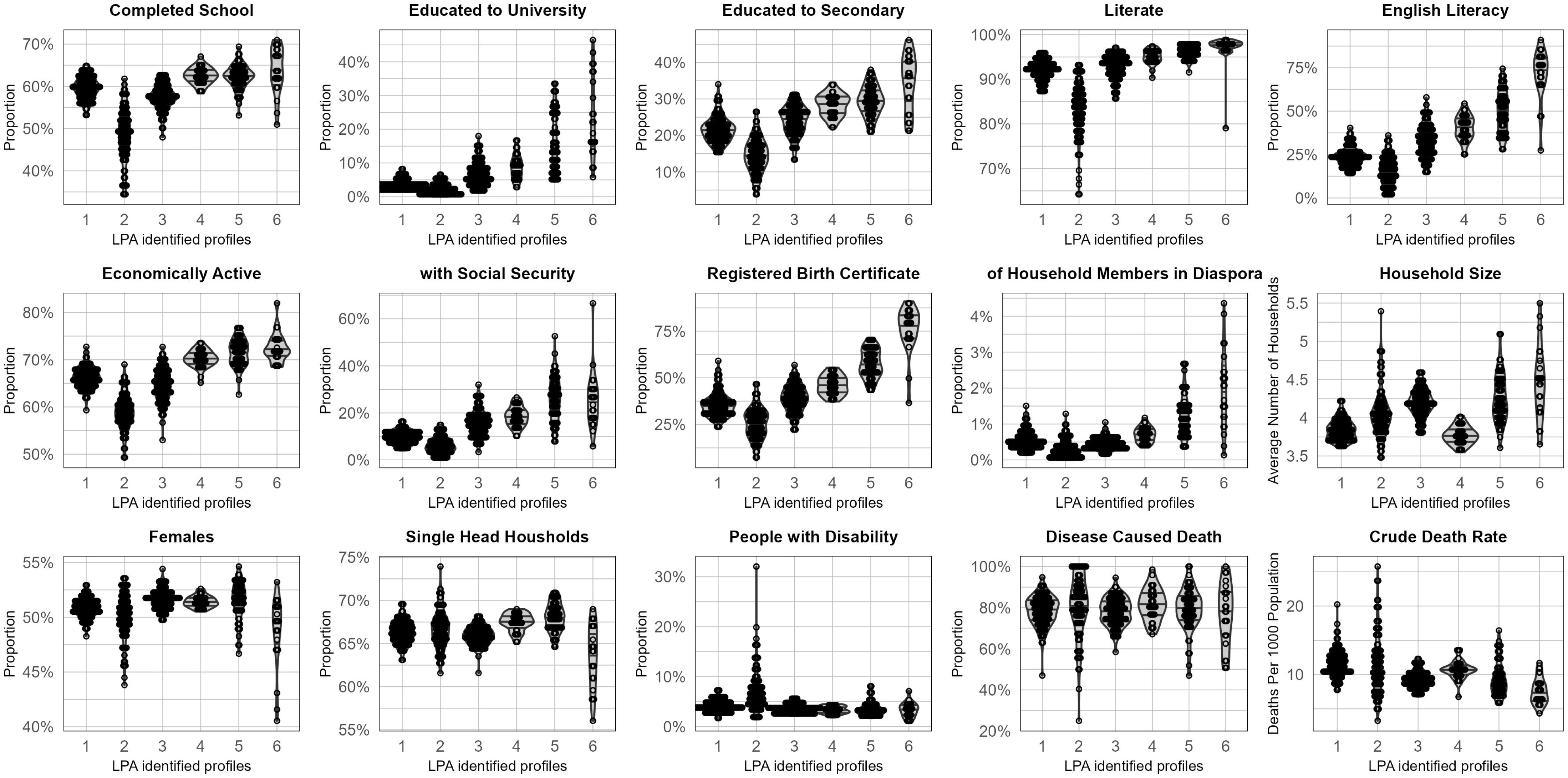
Violin and dot plot showing the distribution of values for 15 potential indicators of deprivation recorded by the 2012 national census, broken down across the six profiles distinguished by LPA analysis (Table 2). Horizontal lines within the violin plot represent the first, second and third quartiles, or the 25%, 50% (median), 75% quantiles. The units for most of these variables are proportions of participating respondents or households, aggregated at neighbourhood level, the only two exceptions being household size and crude death rate.

**Fig 5:**
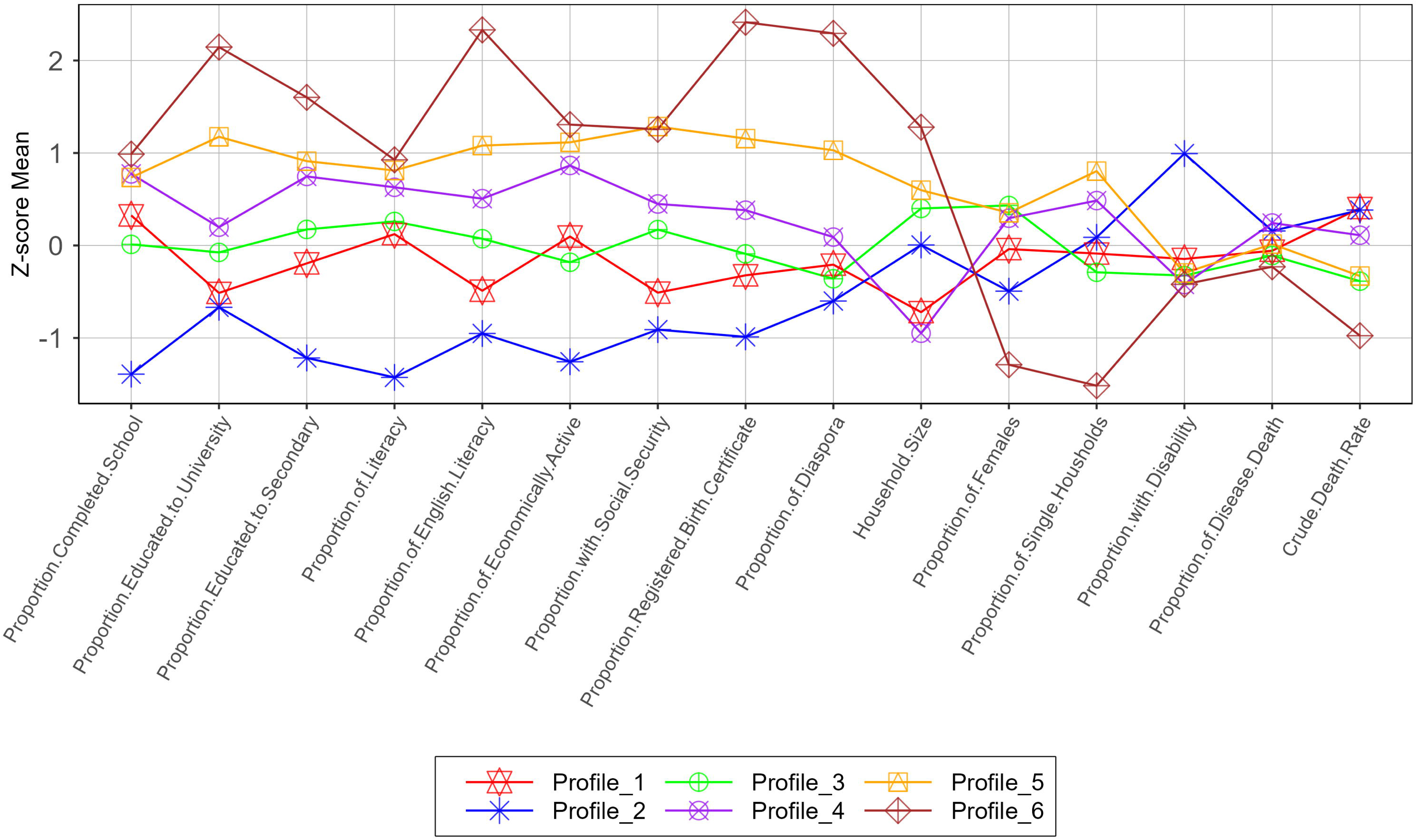
Line and dot plot illustrating the distribution of calculated z-score means for each contributing variable for each profile identified by latent profile analysis (LPA) and subsequently attributed to a named stratum of deprivation (Table 3). Means were scaled by standardizing observed variables (z-scores) within each latent profile and expressing them in standard deviation units instead of the original metric. Scaling transforms the latent variable to have mean of zero and standard deviation of one (μ = 0, σ = 1). Negative means do not imply negative values of the actual metric, but rather reflecting normalization around zero by the scaling process.

**Table 3:**
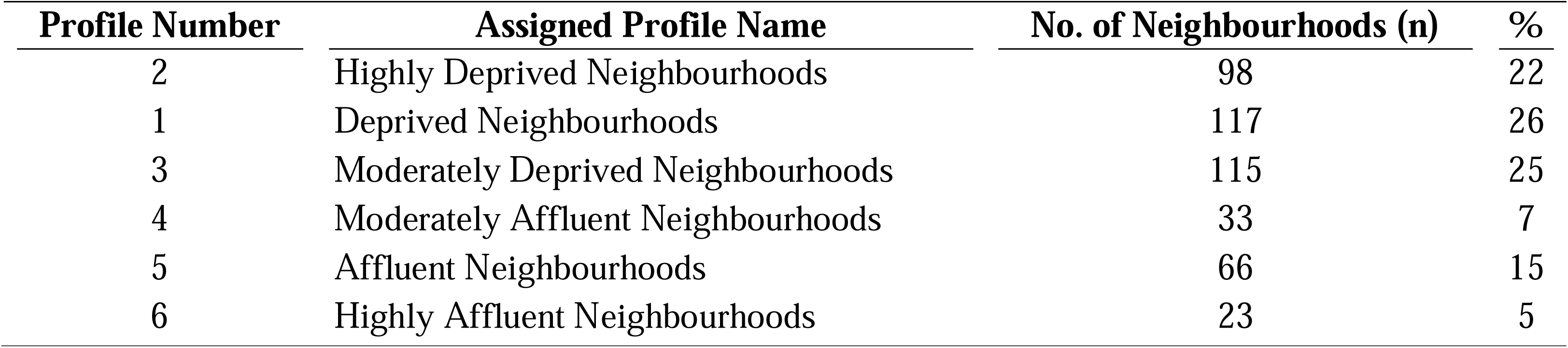
Latent profile analysis (LPA) assigned profiles numbers with investigator assigned ranked deprivation stratum names. Based on interpretation of Fig and Fig 5, with the number and proportion of neighbourhoods attributed to each stratum.

| Profile Number | Assigned Profile Name | No. of Neighbourhoods (n) | % |
| --- | --- | --- | --- |
| 2 | Highly Deprived Neighbourhoods | 98 | 22 |
| 1 | Deprived Neighbourhoods | 117 | 26 |
| 3 | Moderately Deprived Neighbourhoods | 115 | 25 |
| 4 | Moderately Affluent Neighbourhoods | 33 | 7 |
| 5 | Affluent Neighbourhoods | 66 | 15 |
| 6 | Highly Affluent Neighbourhoods | 23 | 5 |

Based on the above interpretation of the LPA identified profiles, six assigned deprivation strata could be resolved from the census data (Table 3). On the other hand, however, the results from participatory stratification approach revealed five income classes (Table 4) with *low income stratum* accounting for a remarkably low percentage of neighbourhoods while the *medium income stratum* had the highest.

**Table 4:** Neighbourhood income classes obtained from participatory stratification approach (PSA), with the number and proportion of neighbourhoods attributed to each stratum.

| Class Number | Assigned Classes Name | No. of Neighbourhoods (n) | % |
| --- | --- | --- | --- |
| 5 | Low Income | 19 | 4 |
| 4 | Mixed Low and Medium Income | 88 | 19 |
| 3 | medium income | 199 | 44 |
| 2 | Mixed Medium and High | 102 | 23 |
| 1 | High Income | 44 | 10 |

### Comparison of PSA with LPA of the census data

The results of the spearman rank correlation test shows that there is a strong positive correlation (ρ = 0.739, S = 4015865, p < 0.001) between the stratum identified by the LPA of census data and the results of the PSA (Fig).

**Fig 6:**
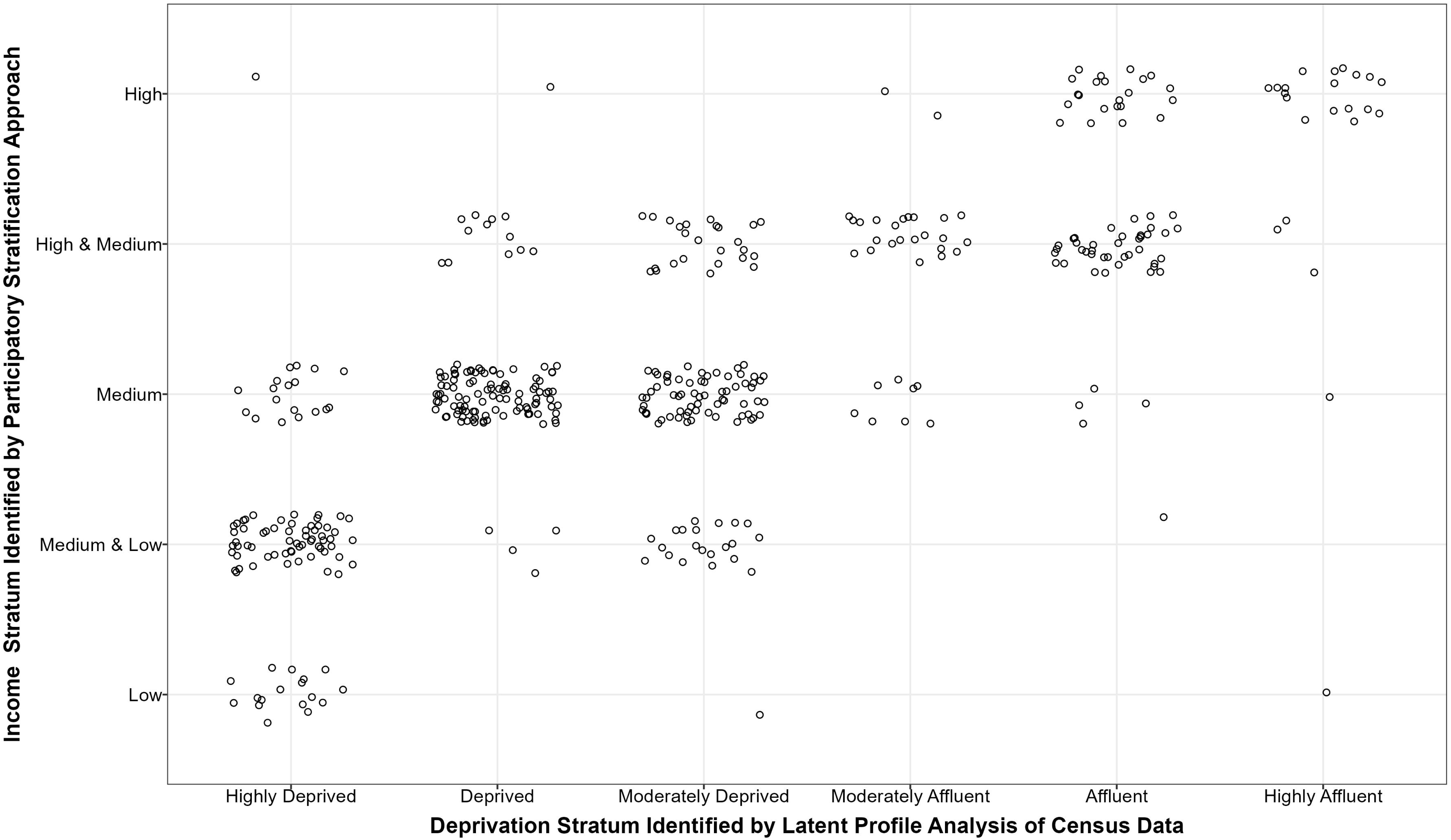
Jitter plot showing the relationship between the neighbourhood classification obtained through the participatory stratification approach (PSA), and through statistical latent profile analysis (LPA) stratification based on national census data.

The same variables used to stratify the neighbourhoods by LPA yielded distribution of values that were just as consistent with their income stratum as determined by the PSA (7) as they were with their deprivation stratum as determined by the LPA (Fig and Fig 5).

**Fig 7:**
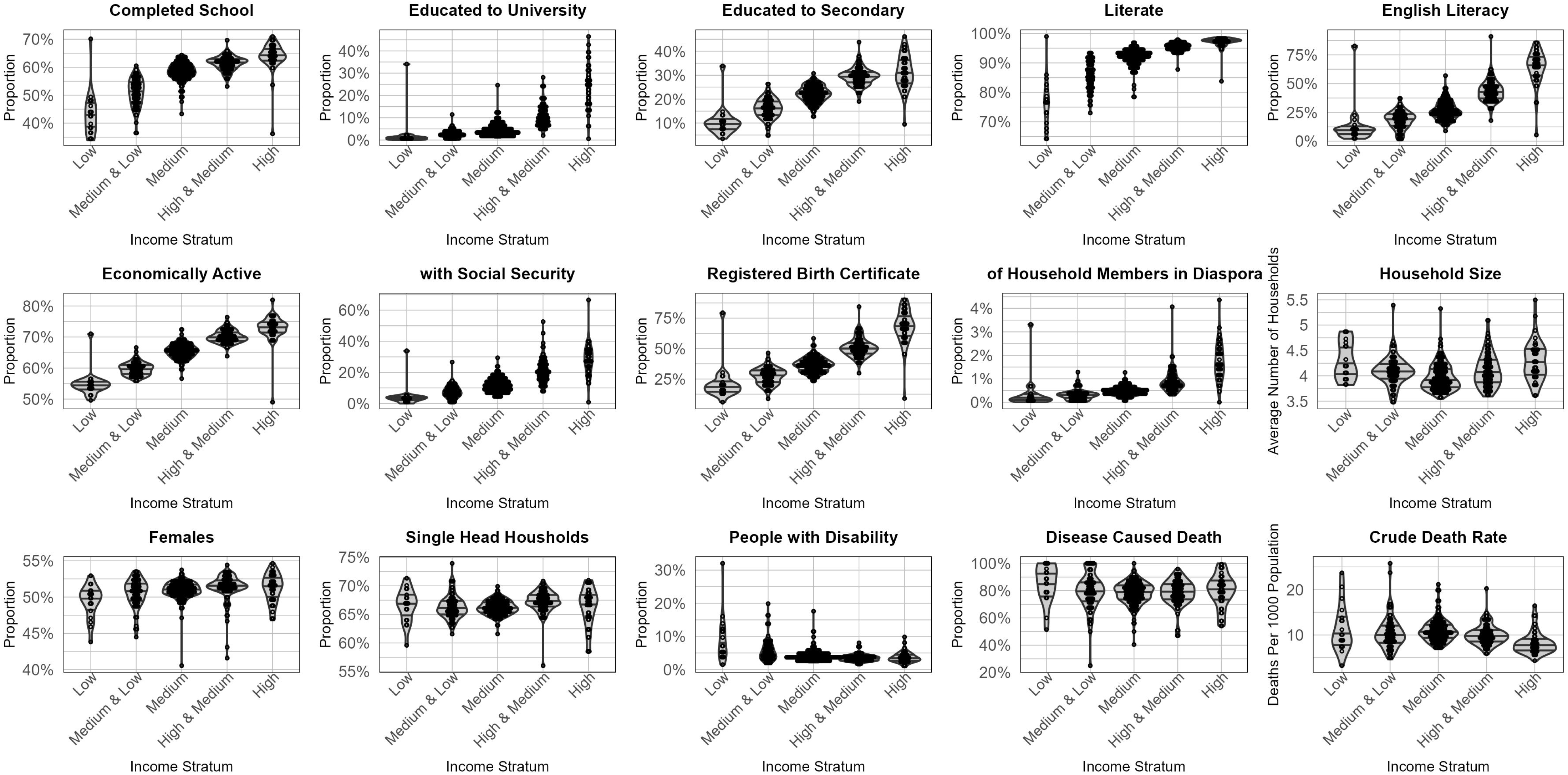
Violin and dot plot showing the distribution of values for 15 potential indicators of deprivation recorded by the 2012 national census, broken down across the five income strata obtained from the participatory stratification approach (PSA). Horizontal lines within the violin plot represent the first, second and third quartiles, or the 25%, 50% (median), and 75% quantiles. The units for most of these variables are proportions of participating respondents’ households aggregated at neighbourhood level, the only two exceptions being household size and crude death rate.

Further analysis of Spearman’s rank correlation tests between each variable from the census data and stratum rank as determined by each of the two different stratification approaches can be seen in 8. A Wilcoxon signed-rank test to compare these two sets of correlation coefficients in a paired manner revealed no difference between the two approaches (V = 33, p-value = 0.1354), indicating that the two approaches yielded very similar patterns of stratification.

**Fig 8:**
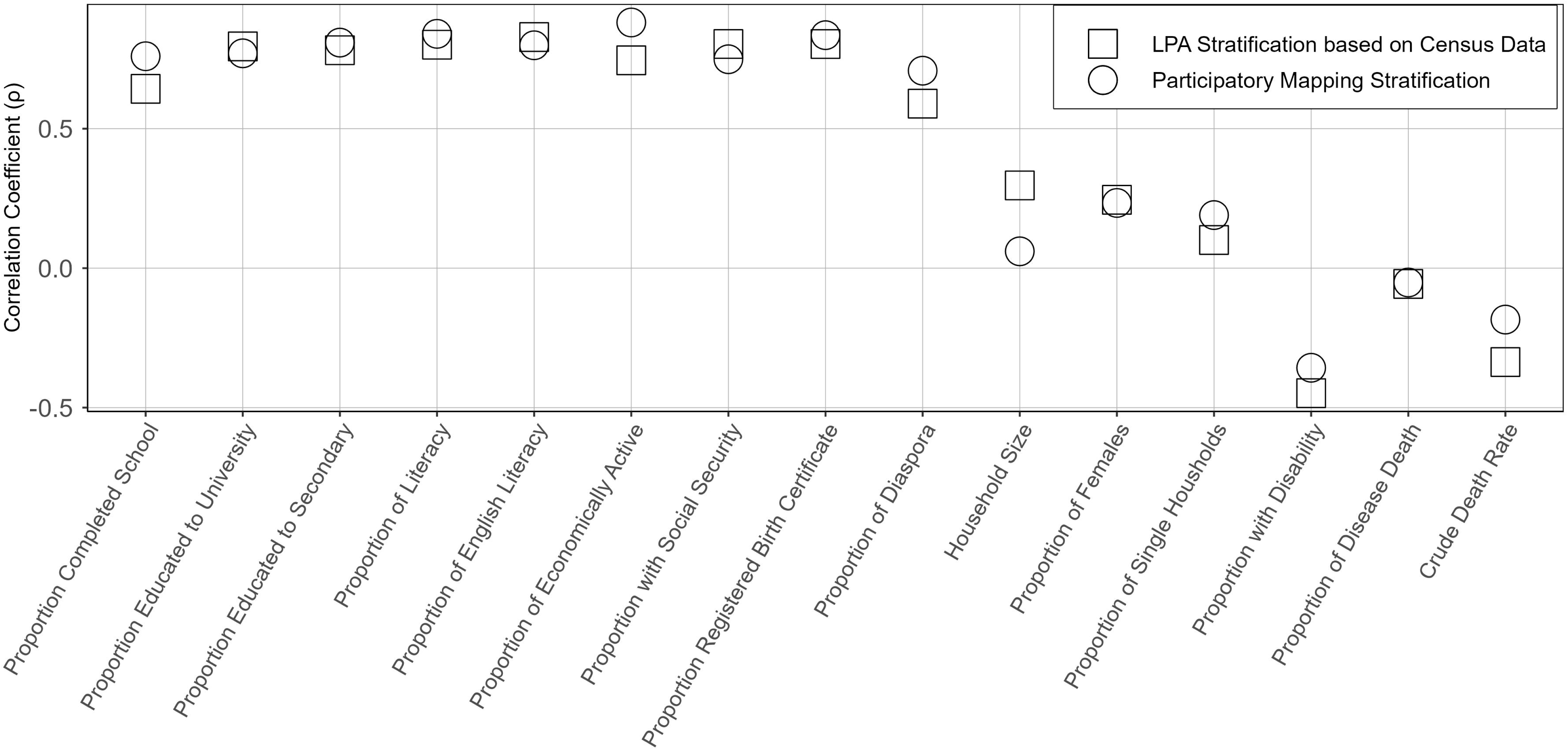
Dot plot showing the results of the Spearman’s rank correlation test of the census variables with each of the two stratification approaches, namely the participatory stratification approach (PSA), and the latent profile analysis (LPA), with vertical axis showing correlation coefficient (ρ) of each variable with the stratum ranking for each stratification method.

Overall, the two stratification approaches yielded relatively similar cartographic pictures of wealth and poverty distribution across the city (Fig). High income, highly affluent neighbourhoods are mainly located in or around the city centre and along the coast to the north, while low income, deprived neighbourhoods are generally distributed along the outskirts of the city. The distribution pattern of the medium income stratum, as determined by PSA, corresponds well with the moderately deprived and moderately affluent strata identified by LPA. Focusing on the city centre and areas immediately around it, the results reveal patches of low income and deprived neighbourhoods located near high income and highly affluent neighbourhoods, forming a pocket of poverty running from central Ilala into Temeke District. However, in the western direction this pocket of deprived, medium income neighbourhoods is enclosed by moderately deprived, mixed medium and high-income neighbourhoods.

The overall distribution, on the other hand, shows that most of the neighbourhoods classified by PSA as medium income in Fig A, such as Msimbazi Bondeni and Kigogo Kati, havebeen classified as deprived neighbourhoods by LPA in Fig B. Many of these neighbourhoods are found in the same pocket of deprived neighbourhoods described above that cuts across all three districts but mostly sits in northern Temeke.

**Fig 9:**
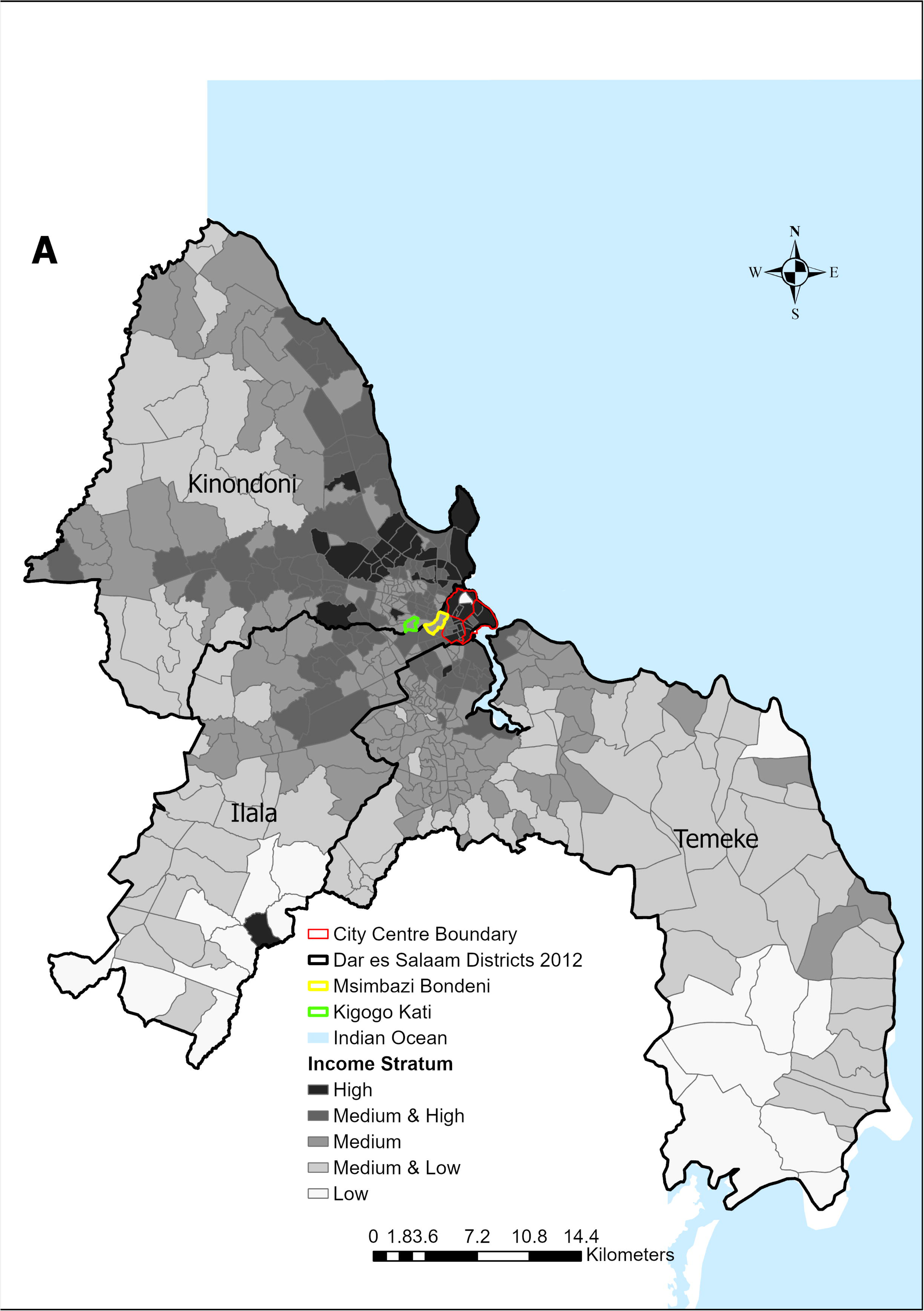
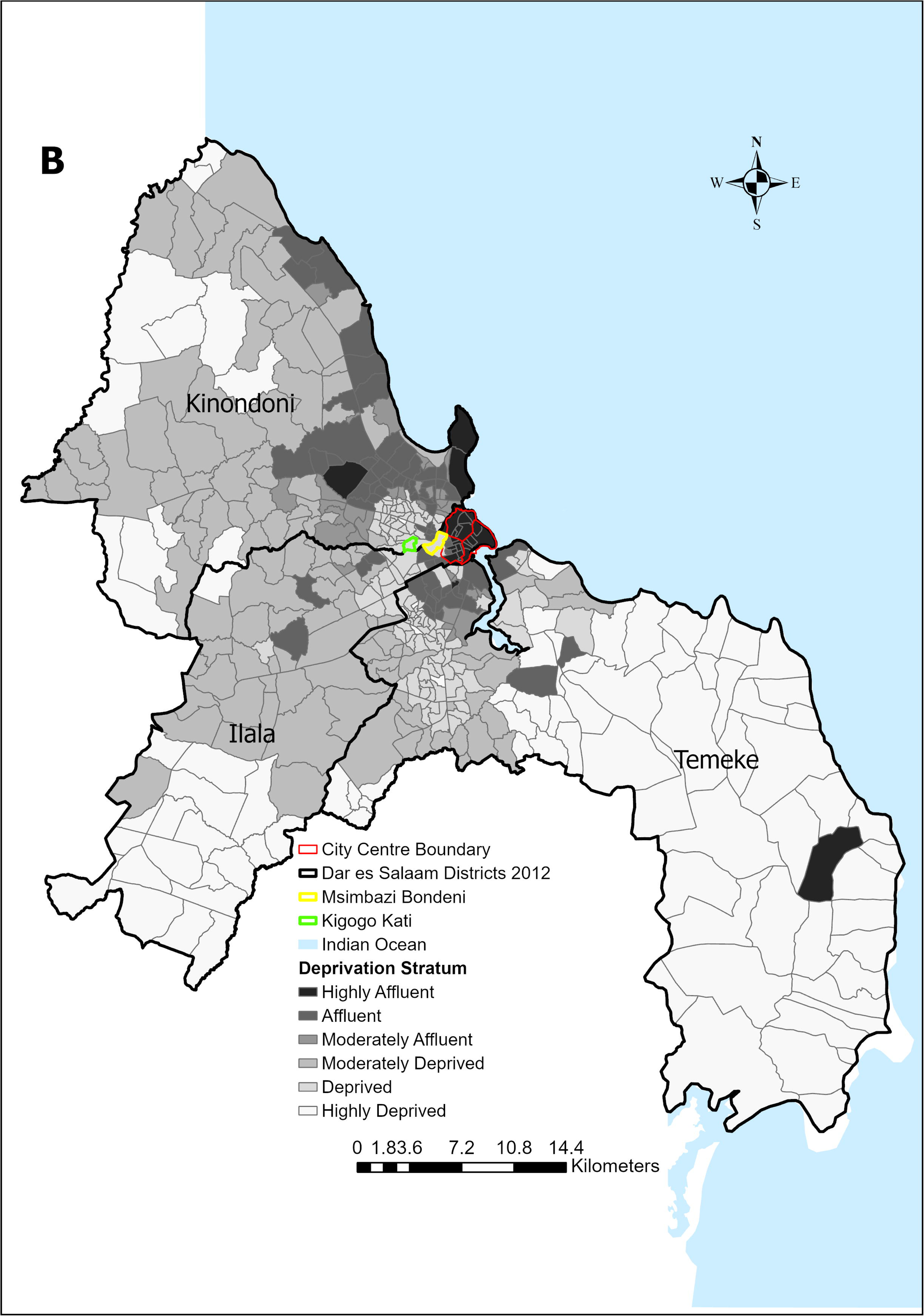
The spatial distribution of income and deprivation strata across Dar es Salaam city neighbourhoods. Panel A: Income strata obtained from the participatory stratification approach (PSA) (Table 4). Panel B: deprivation strata generated from a statistical latent profile analysis (LPA) (Table 3) of population census data.

Looking at the heterogeneous settlement patterns of these neighbourhoods sheds light on how they were classified differently by the two different methods (Fig and Fig). For example, the varying density and quality of the buildings in Kigogo Kati illustrated in Fig shows that, while participants in the PSA stratification process considered the mixed housing qualities and density that are clearly visible in panel A to be a reasonable basis for classifying it as medium income, LPA averaged out the diversity existing at fine scale within the neighbourhood and consequently classified it simply as being deprived.

Additionally, while the example in Fig outlines the spatial distribution of different building qualities within the same neighbourhood, similar variability is clearly noticeable within some other neighbourhoods. For example, Fig illustrates part of the Msimbazi Bondeni neighbourhood, where two housing blocks with obviously different building qualities are found in the same neighbourhood, which has been classified as medium income by PSA but deprived by LPA. Moreover, these two blocks with obviously different mean housing density and quality lie at opposite sides of a steep topographic incline (Fig A and B), with the denser informal settlements with generally smaller, poor quality housing being found in the flood-prone valley to the north of it, while the carefully constructed apartment blocks and planned housing developments (Fig D and E) were all found approximately 20 meters uphill to the south. Indeed, as one walks southward along the transect depicted in Fig 7A and B, from the valley up the hill onward along a relatively flat plateau, a clearly noticeable shift is observed in building quality with elevation (Fig, panel D and E).

Although there are some differences between the two approaches in the distribution patterns of income and deprivation across neighbourhoods, with very few being classified as purely low income by PSA (Table 4), the geographic distribution of poverty displayed by the two approaches (Table 3 and Table 4) nevertheless follow a similar pattern (Fig).

## Discussion

This study contributes to a growing area of research demonstrating similarities and differences in the results obtained through quite different methodological approaches to neighbourhood stratification. The use of LPA has provided valuable insights into the classification of neighbourhoods, particularly in the assessment of deprivation profiles. The results demonstrate LPA’s ability to identify statistically distinct discrete neighbourhood deprivation profiles using socioeconomic data, while highlighting its limitation in missing out the complex realities existing at a finer spatial scales within neighbourhoods. This, however, is just one facet of the broader challenge posed by profiling deprivation using aggregated data at the level of the neighbourhoods in which people live, especially in diversified cities like Dar es Salaam. Furthermore, through cross-validation of the two approaches, both statistically and spatially, the results of the PSA yielded very similar patterns of heterogeneity as the LPA, in terms of the distributions of variable values and neighbourhood classifications, respectively. Furthermore, PSA proved capable of elucidating several hidden factors relying on perceived and lived understanding of poverty, which were easily missed by LPA (**Fig** 0 &**Fig** 1). Akin to many participatory approaches [17, 20, 45–49] relying on the subjective perspectives of contributing participants, the PSA developed in this study, proved to be cost-effective and user-friendly. In contrast, while statistical analyses of formally collected data, such as the LPA approach described herein, have the advantage of being objective, they are also time-consuming, data-intensive and require advanced analytical skills.

Nevertheless, the subjectivity of participatory approaches has been confirmed by the numbers of neighbourhoods attributed to each income stratum. While PSA classified only 4% of the city’s neighbourhoods as typically low-income (Table 4), LPA classified 22% as highly deprived neighbourhoods (**Table 3**). On the other hand, PSA classified 44% of the city’s neighbourhoods as medium income, indicating a bias away from extreme classification assignments. The surprisingly low frequency with which neighbourhoods were classified as low income by PSA participants implies the subjectivity of the approach, as even some small pockets of better housing in a neighbourhood known to be generally very poor often prompted participants to classify it as medium income.

Furthermore, these PSA results seem incompatible with those of previous studies indicating widespread that large African cities accommodate large deprived populations[50, 51]..

Additionally, the observation that PSA classified some of the neighbourhoods as medium income while LPA classified them as deprived, requires consideration of the fundamentally different strengths and weakness of these two complementary approaches. A possible explanation for this apparent bias of PSA towards moderate stratum assignment may well lie in the diversity existing at fine scales within neighbourhoods. Such fine-scale heterogeneities can easily be missed by the objective statistical approaches but captured, and perhaps subjectively over-represented, by the qualitative understanding of the city obtained through the lived experiences of the PSA participants, wherein even very small differences in housing quality and infrastructures within neighbourhoods were considered in the stratification. For example, the small blocks of high quality, low density housing that are highlighted in yellow bordered panels in Figs 10 and 11 resulted in these neighbourhoods being classified as medium income, despite being well known as poor neighbourhoods (Fig and Fig). Also, the confirmatory stage of the consensus building process in the PSA workshop, resulted in participants providing supplementary justification for their initial income stratum assignment, taking into consideration additional factors that varied across coarser scales than income *per se*, such as physical accessibility, closeness to the valley, road condition, roof age and building density. These intuitive considerations, and ability of the PSA to accommodate them in a flexible manner, differentiates the fine-scale lens of PSA from the coarser analysis of the LPA, which unavoidably blurs such heterogeneities within neighbourhoods.

**Fig 10:**
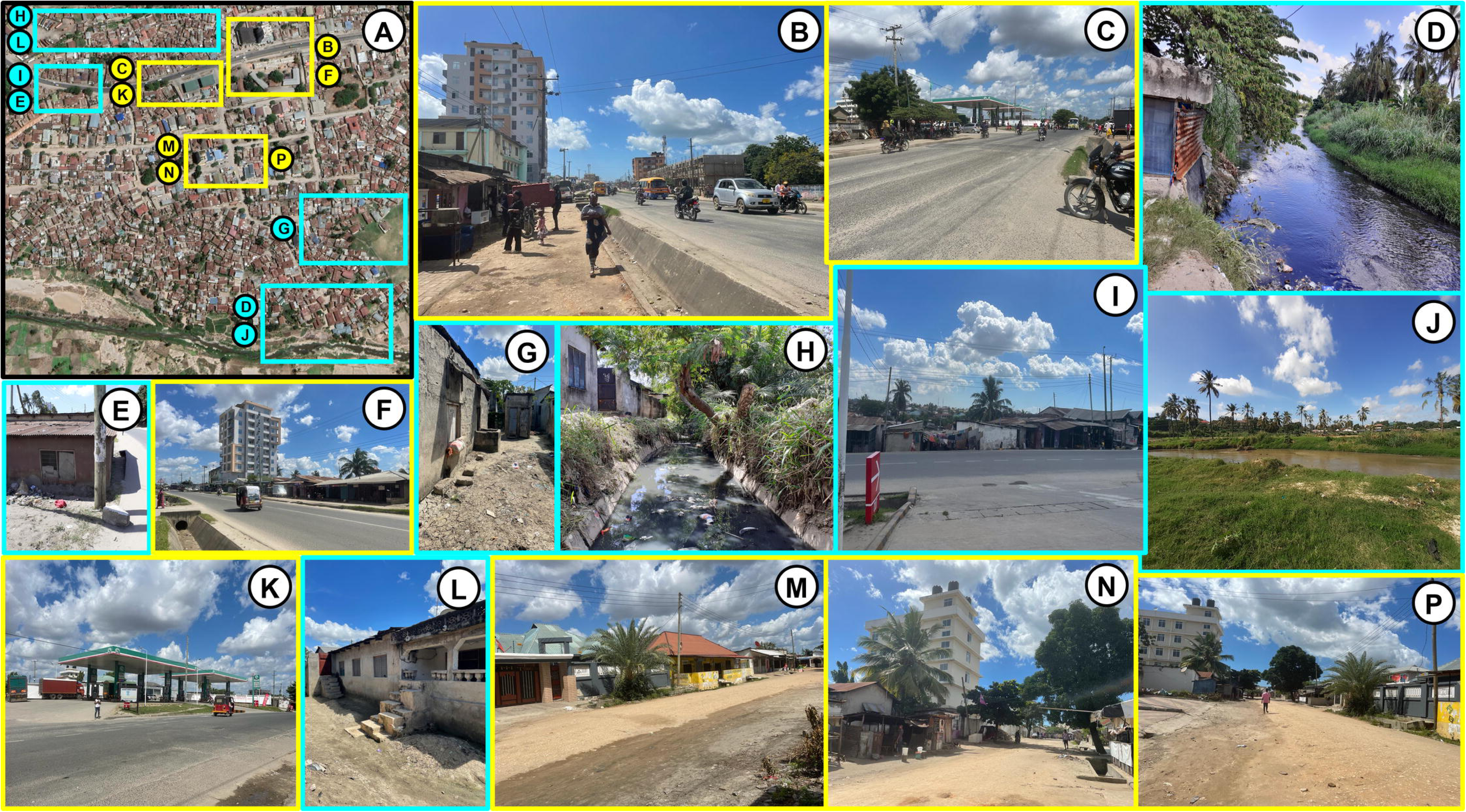
Satellite and building images from the Kigogo Kati neighbourhood showing fine-scale variations in housing density and building quality within the neighbourhood. Kigogo Kati was classified as medium income by participatory stratification approach (PSA) (Fig A), but as deprived by statistical latent profile analysis (LPA) of national census data (Fig B). As mapped out in panel A, blocks of high-quality buildings (yellow bordered panels, B, C, F, K, M, N and P), are contrasted with blocks with generally poor quality housing (blue bordered panels D, E, G, H, I, J and L) in the same neighbourhood.

**Fig 11:**
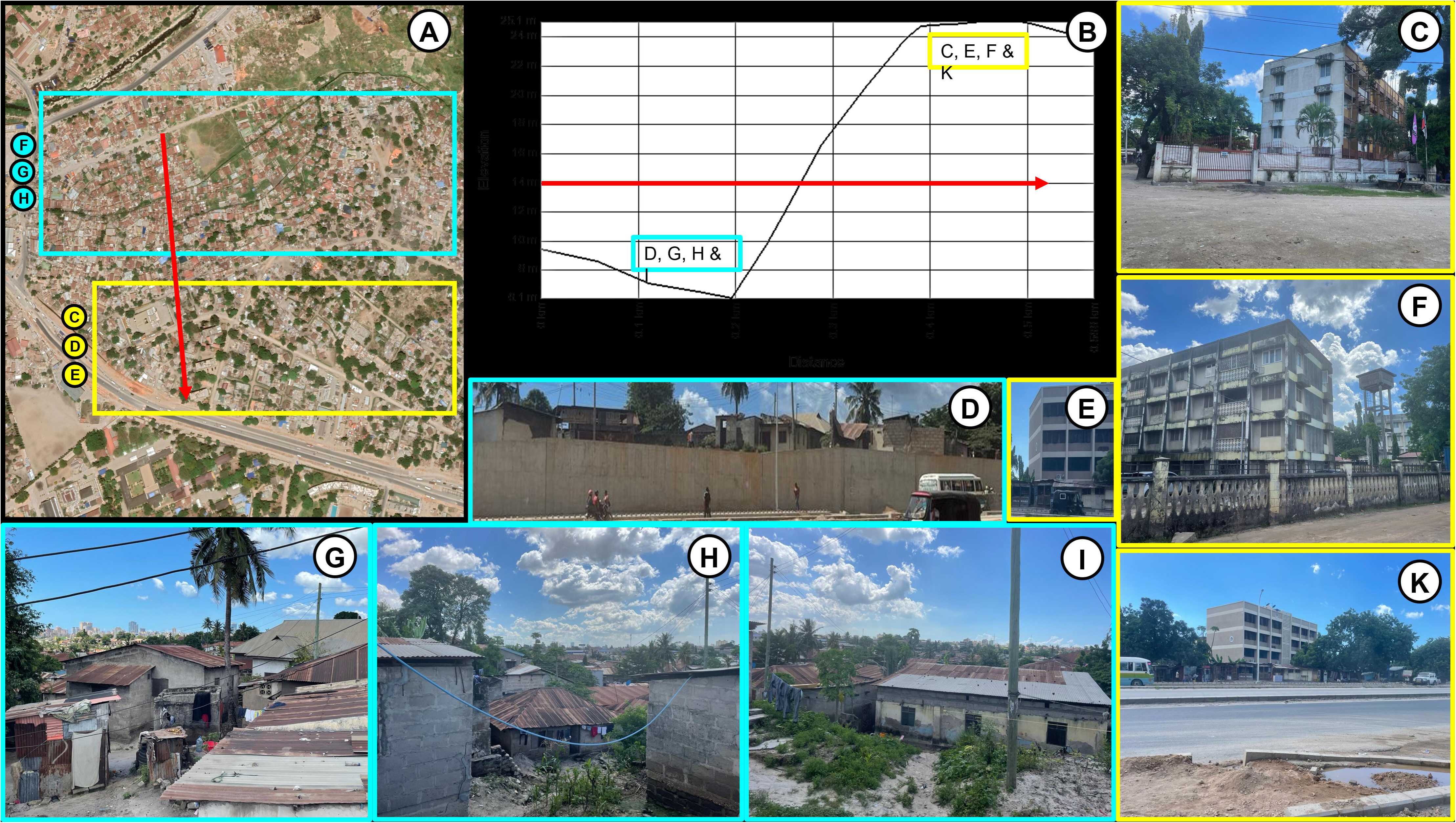
Satellite and building images from the Msimbazi Bondeni neighbourhood, showing fine scale variation in housing density and building quality within the neighbourhood. Msimbazi Bondeni was classified as medium income by participatory stratification approach (PSA) (Fig A) but as deprived by statistical latent profile analysis (LPA) of national census data (Fig B). As mapped out in Panel A, blocks of high-quality buildings (yellow bordered panels, C, E, F and K) uphill and to the south of a steep topographic gradient (Panel B) are contrasted with downhill blocks to the south where high-density settlements have generally poor-quality housing (blue bordered panels D, G, H and I). Panels A and B illustrate how building quality changes with elevation, with the vertical axis in panel B projecting elevation above sea level, while the horizontal axis represents the distance southward along the red vector in panel A.

Such mixed contexts at fine scales might also help explain some of the more puzzling outliers in Fig, because the nuanced, intuitive and holistic basis of the PSA could result in it being subjectively influenced by the most obvious features and characteristics of a neighbourhood in a disproportionate manner, whereas the LPA relies on an objective representation of the area as a whole. Nevertheless, the two approaches yielded statistically equivalent results, even though an intuitive conceptual rationale was used in selecting the quantitative deprivation indicators used for LPA, while qualitative participants knowledge were used in PSA. Hence, the two alternative approach can be applied to complement each other and in different scenarios with different application requirements for stratifying neighbourhoods [52].

This study study contributes to the growing area of research by demonstrating methodological similarity and differences existing in neighbourhood stratification. It complements the existing literature on economic inequality and urban segregation [33, 53–59], which relies for the most part on statistical analysis of empirical data. Furthermore, it suggests that this more subjective participatory approach, which exploits community knowledge of the location, lived experiences and local expertise, could be used in the future without any such data-intensive statistical approaches to validate it. Overall, this study provides clear evidence that cost-effective PSA methodologies could have a variety of practical applications, such as services planning and local tailoring of urban interventions.

While this research reassuringly presents neighbourhood-level income and deprivation profiles, respectively assessed through a subjective PSA and an objective LPA of formal socioeconomic data, some important study limitations are worth mentioning. For example, the certainty of the findings may be somewhat limited by the fact that, at the time of this study, there were no more recent census data available than those from the 2012 PHC, so the results obtained with the two methods cannot be considered exactly contemporaneous. Similarly, the retrospective *post hoc* examination of heterogeneities within neighbourhoods (**Fig** and **Fig**) relies on images of contemporary Dar es Salaam, which has obviously changed somewhat since the time that the PSA was conducted and the PHC data used for LPA were collected.

Moreover, the absence of data relating to variables like employment, household incomes, neighbourhood incomes, property ownership, and living conditions (including housing, water, and electricity) restricted our ability to establish a conclusive relationship between income and living standards on one hand and deprivation on the other. These factors are traditionally utilized as indicators for assessing deprivation across various geographical and poverty dimensions. Consequently, future studies should explore alternative poverty indicators and incorporate updated data when accessible.

Additionally, the study used area-based data aggregated at neighbourhood level, which was the finest administrative scale made available to the investigators, so the stark heterogeneities that clearly exist within individual neighbourhoods (**Fig** 0 and **Fig** 1), are obviously blurred around the neighbourhood average income by the LPA and cause underrepresentation of the lowest income neighbourhoods by PSA (**Fig**). Correspondingly, further research is needed to similarly compare subjective PSAs with objective statistical stratification based on formal survey data at finer scales, which in the context of Dar es Salaam, for example, could relate to ten cell unit housing clusters. Other avenues for future research can also include studies on how the built environment influences income and deprivation, and how the physical geography of the landscapes those human settlements are built upon affects patterns of income, access to services, health and wellbeing among residents.

## Conclusion

The purpose of this study was to evaluate a PSA to stratifying neighbourhoods across the Tanzanian city of Dar es Salaam in terms of income, by comparing with LPA of deprivation based on national census data. The findings revealed the potential of both LPA and PSA methods and set a precedent for their further application as complementary approaches to neighbourhood classification. With reference to PSA, this study highlights ample room for advancing our understanding of the complex relationships between poverty, location, and socio-spatial dynamics in urban settings. To achieve a more comprehensive perspective, we recommend moving beyond reliance on relatively coarse geographic units for aggregating large-scale datasets. Instead, we propose adopting a finer-grained approach that can capture previously overlooked social, economic, physical, and environmental factors at the very fine scales they can vary across. Overall, such methodologies may be valuable, not only for policy makers, but also for urban planning practitioners and researchers who seek a deeper understanding of the diverse dynamics occurring within towns and cities across various scales. These findings confirm that subjective PSA methodologies offer more affordable and practical alternatives to conventional objective statistical analyses of formal survey data, with which to obtain detailed geographic information to inform decision-making and planning in towns and cities all across low and middle-income countries.

## Data Availability

The data underlying the results presented in the study are available from the National Bureau of Statistics of Tanzania https://www.nbs.go.tz/index.php/en/

## Acknowledgements

We gratefully acknowledge the support and contributions that made this research possible. Special thanks to Prof. Ya Ping Wang and Keith Kintrea for their invaluable advice. We also thank Dr. Victoria Mwakalinga for leading the study participants team and all participants from DFID, DAWASA, Ardhi University, Humanitarian Open street Team, Ifakara Health Institute, Ministry of Lands Housing and Human Settlements, Regional Administrative Offices, TARURA, Twaweza and all five Dar es Salaam municipalities by then for their participation in the neighbourhood stratification. This work was made possible by the collective efforts of all those mentioned, and we appreciate their essential roles in the successful completion of this study.

## Supporting information

**S1 Table: Equation used to derive census data and transform selected variables into proportion for each sample neighbourhood.**

**S2 Fig: Pearson correlation plot with correlation coefficients and p-values, showing numerical correlation coefficient labels only for pairs which are strongly correlated (**ρ ≥ **+/- 0.4) and statistically significant (p** ≤ **0.05) while insignificant variables are unlabelled.**

**S3 Fig: Pairwise scatter plots of all the variables as well as histograms, locally smoothed regressions, and Pairwise Pearson’s correlation test matrix with correlation coefficients (**ρ**) and p values (* p < 0.05, ** p < 0.01, *** p < 0.001). The x axis in each scatter plot represents the column variable, the y axis the row variable. The units for most of these variables are proportions of participating respondents or households, aggregated at neighbourhood level, the only two exceptions being household size and crude death rate.**

---

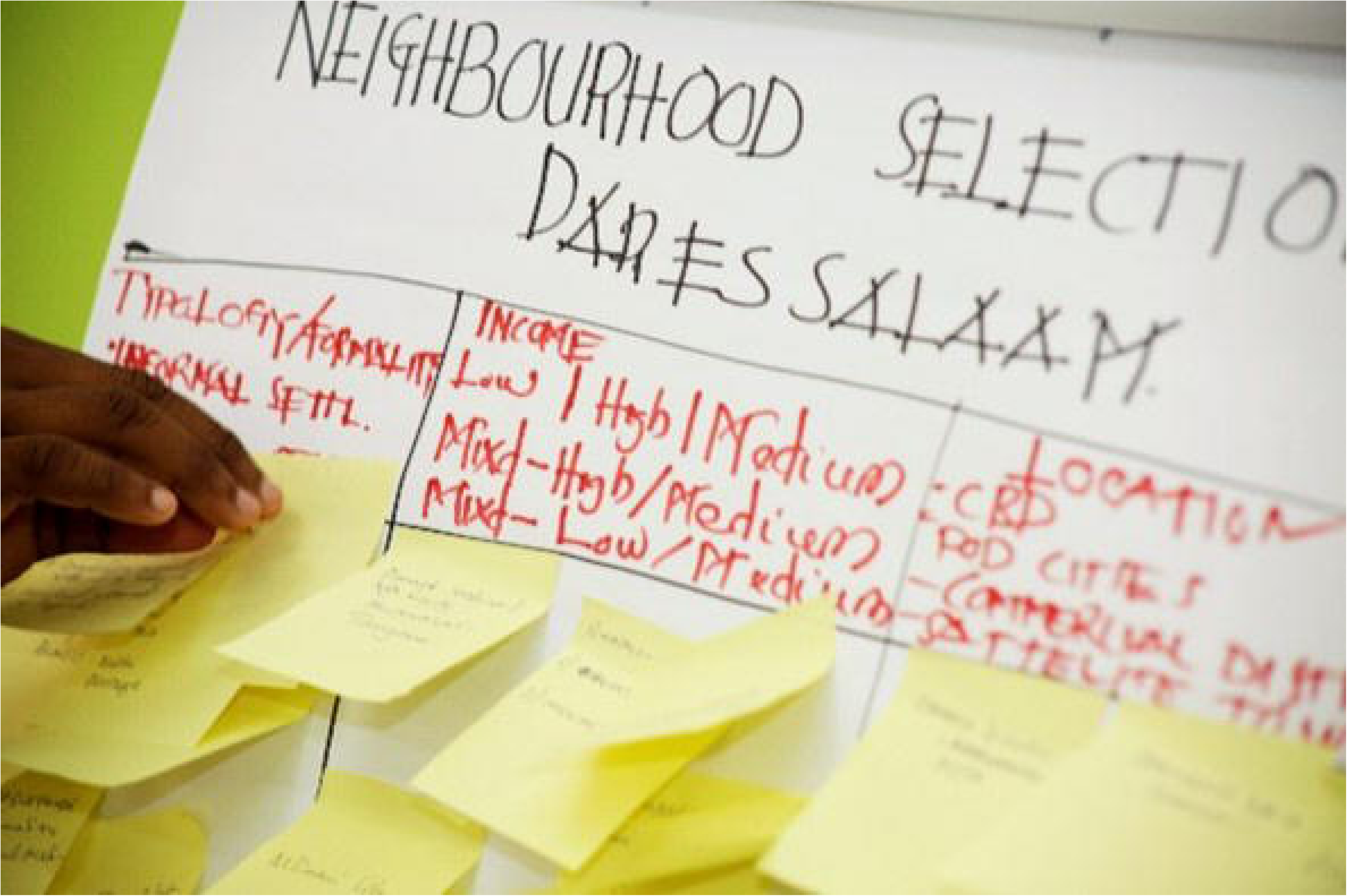

## Notes

### Competing Interest Statement

The authors have declared no competing interest.

### Funding Statement

This work was conducted as part of the Centre for Sustainable Healthy and Learning Cities and Neighbourhoods funded via UK Research and Innovation and administered through the Economic and Social Research Council as part of the UK Governments Global Challenges Research Fund. Project Reference: ES/P011020/1.
The funders had no role in study design data collection and analysis decision to publish or preparation of the manuscript.

### Author Declarations

Ethical clearance for the study, a component of the Sustainable Cities project at Ifakara Health Institute (IHI), was granted by the Institutional Review Board of the Ifakara Health Institute (IHI/IRB/NO: 26-2018) and the Medical Research Coordinating Committee at the National Institute for Medical Research in Tanzania (NIMR/HQ/R.8a/Vol. IX/2954).

