## Supporting Table 1, Supporting Figure 2 and 3 for "Evaluation of a participatory approach to stratifying neighbourhoods across the Tanzanian city of Dar es Salaam in terms of income by comparing with latent profile analysis of deprivation based on national census data": S1 Table.docx

| **Category** | **Variable** | **Equation** |
| --- | --- | --- |
| Education | Proportion completed any school level | $=\left( \frac{CompleSch}{(CompleSch+AttendingSch+PartlyAttended+NeverAttended)} \right)$ |
|  | Proportion educated to university level | $=\left( \frac{UniversityOther}{(NurseryEdu+Primary\left\{ 1,8 \right\}+TrainingAfterPri+PreFormOne+Secondary\left\{ 1,6 \right\}+TrainingAfterSec+UniversityOther)} \right)$ |
|  | Proportion educated to secondary level | $=\left( \frac{Secondary\left\{ 1,6 \right\}}{(NurseryEdu+Primary\left\{ 1,8 \right\}+TrainingAfterPri+PreFormOne+Secondary\left\{ 1,6 \right\}+TrainingAfterSec+UniversityOther)} \right)$ |
| Literacy and language proficiency | Proportion of literacy | $=\left( \frac{(Total Population-Illiterate)}{Total Population total} \right)$ |
|  | Proportion of English literacy | $=\left( \frac{\left( English+SwahiliEnglish \right)}{(Swahili+English+SwahiliEnglish+Othelanguage)} \right)$ |
| Economic factors | Proportion of economically active individuals | $=\left( \frac{N\mathrm{age}\left\{ 15,64 \right\}}{Nage\left\{ 0,80+ \right\}} \right)$ |
|  | Proportion with social security | $=\left( \frac{socialsec}{(socialsec+nosocialsec)} \right)$ |
| Social and legal indicators | Proportion registered birth certificate | $=\left( \frac{BirthCertificate}{(BirthCertificate+BirthNotification+noCertNot+bircernotknow)} \right)$ |
| Economic factors | Proportion household members in diaspora | $=\left( \frac{Diaspora}{Total Population} \right)$ |
| Demographic factors | Average household size | $=\left( \frac{Total Population}{Total Household Heads} \right)$ |
|  | Proportion of female in a neighbourhood | $=\left( \frac{Females}{Total Population} \right)$ |
|  | Proportion of single head households | $=\left( \frac{\left( NeverMarried+Divorced+Separated\&+Widowed \right)}{\left( NeverMarried+Married+LivingTogether\&+Divorced+Separated+Widowed \right)} \right)$ |
| Health and mortality statistics | Proportion with disability | $=\left( \frac{(Albino+Seeing+Hearing+Walking+Remembering+SelfCare+otherDisability)}{Total Population} \right)$ |
|  | Proportion of death caused by diseases | $=\left( \frac{Total Disease Death}{Total Death} \right)$ |
|  | Crude death rate | $=\left( 1000*\left( \frac{Total Death}{Total Population} \right) \right)$ |
