## Supplementary figures and images for "Evaluation of a participatory approach to stratifying neighbourhoods across the Tanzanian city of Dar es Salaam in terms of income by comparing with latent profile analysis of deprivation based on national census data"

### S2 Fig.tiff

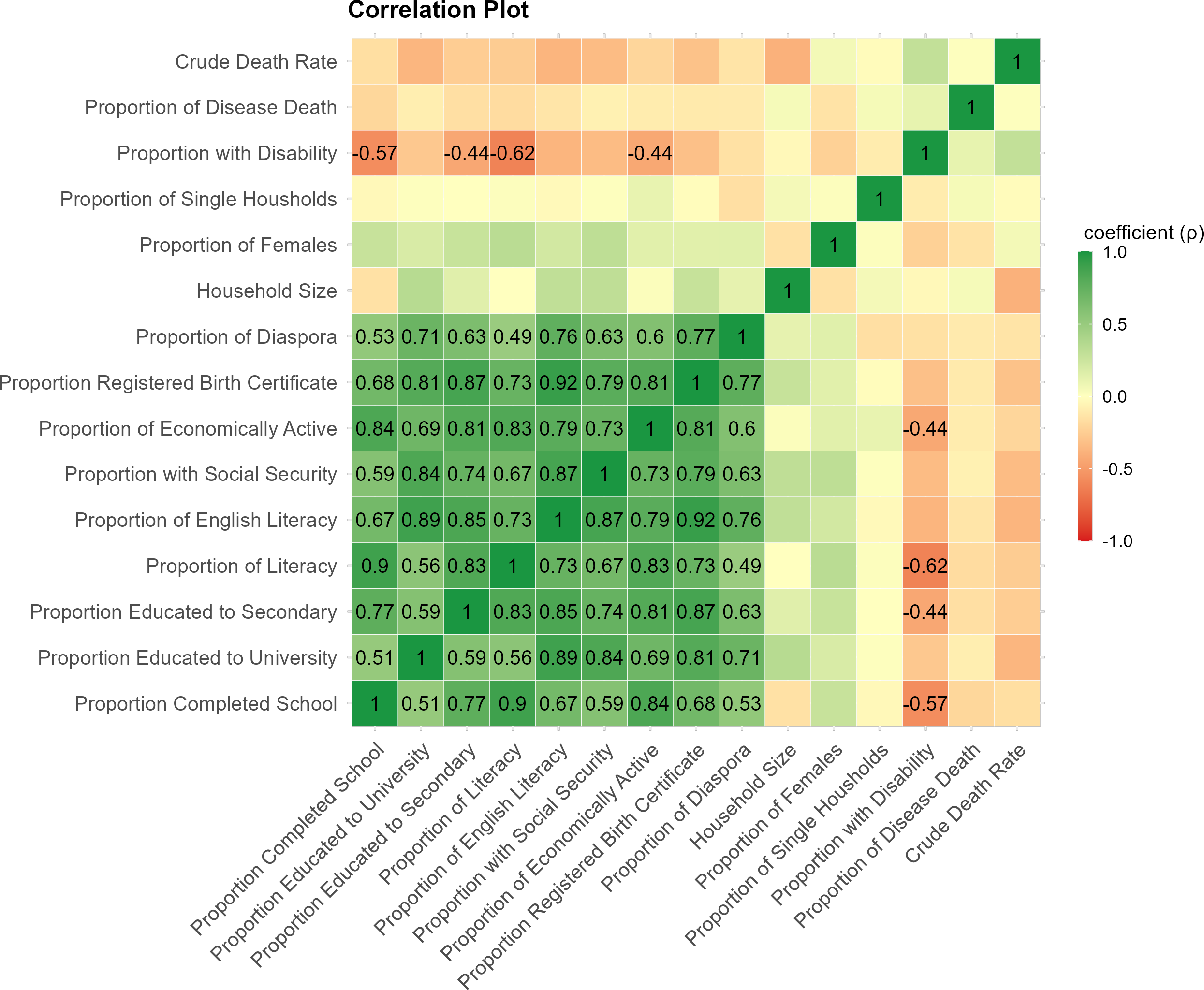

### S3 Fig.tiff

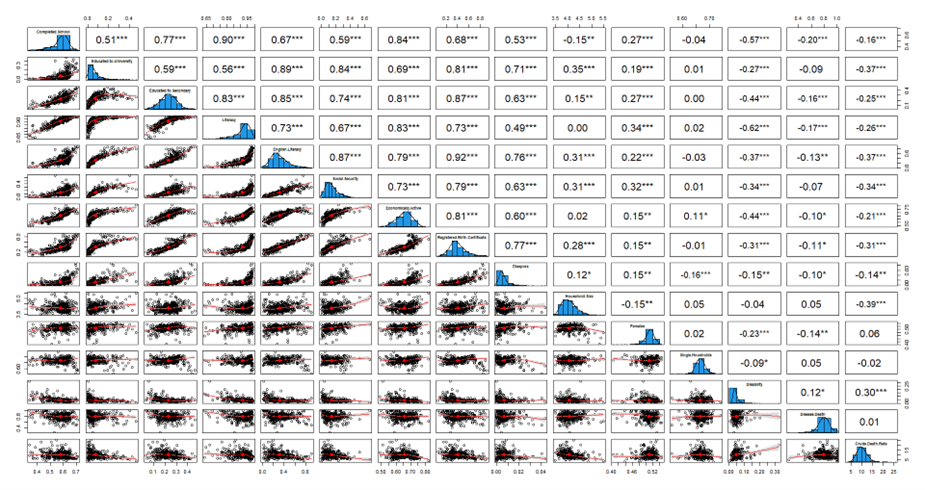
